# Massachusetts General Hospital Covid-19 Registry reveals two distinct populations of hospitalized patients by race and ethnicity

**DOI:** 10.1101/2020.09.08.20190421

**Authors:** Ingrid V. Bassett, Virginia A. Triant, Bridget A. Bunda, Caitlin A. Selvaggi, Daniel J. Shinnick, Wei He, Frances Lu, Bianca C. Porneala, Tingyi Cao, Steven A. Lubitz, James B. Meigs, John Hsu, Andrea S. Foulkes

## Abstract

**Objective:** To evaluate differences by race/ethnicity in clinical characteristics and outcomes among hospitalized patients with Covid-19 at Massachusetts General Hospital (MGH).

**Methods:** The MGH Covid-19 Registry includes confirmed SARS-CoV-2-infected patients hospitalized at MGH and is based on manual chart reviews and data extraction from electronic health records (EHRs). We evaluated differences between White/Non-Hispanic and Hispanic patients in demographics, complications and 14-day outcomes among the N=866 patients hospitalized with Covid-19 from March 11, 2020 – May 4, 2020.

**Results:** Overall, 43% of patients hospitalized with Covid-19 were women, median age was 60.4 [IQR=(48.2, 75)], 11.3% were Black/non-Hispanic and 35.2% were Hispanic. Hispanic patients, representing 35.2% of patients, were younger than White/non-Hispanic patients [median age 51y; IQR=(40.6, 61.6) versus 72y; (58.0, 81.7) (p< 0.001)]. Hispanic patients were symptomatic longer before presenting to care (median 5 vs 3d, p=0.039) but were more likely to be sent home with self-quarantine than be admitted to hospital (29% vs 16%, p< 0.001). Hispanic patients had fewer comorbidities yet comparable rates of ICU or death (34% vs 36%). Nonetheless, a greater proportion of Hispanic patients recovered by 14 days after presentation (62% vs 45%, p< 0.001; OR=1.99, p=0.011 in multivariable adjusted model) and fewer died (2% versus 18%, p< 0.001).

**Conclusions:** Hospitalized Hispanic patients were younger and had fewer comorbidities compared to White/non-Hispanic patients; despite comparable rates of ICU care or death, a greater proportion recovered. These results have implications for public health policy and the design and conduct of clinical trials.

## Introduction

Massachusetts has over 110,000 cumulative confirmed Covid-19 cases and over 8000 deaths as of August 2, 2020.^1^ The Massachusetts epidemic has a high degree of geographic and racial/ethnic heterogeneity in reported cases, even among cities with similar population density.^2^
 The disproportionate burden of Covid-19 borne by some communities has been attributed to higher proportion of essential workers in public facing jobs, greater household size, and poverty limiting access to care.^
3–5
^ Our objective was to compare clinical and demographic characteristics, complications and 14-day outcomes among Covid-19 patients hospitalized at Massachusetts General Hospital (MGH) by race and ethnicity. MGH is a quaternary care facility with the largest number of hospitalized and intensive care unit (ICU) patients with Covid-19 in the state.

## Methods

The MGH Covid-19 Data Registry includes confirmed SARS-CoV-2-infected patients hospitalized at MGH and is based on manual chart reviews and data extraction from electronic health records (EHRs). Trained reviewers collected demographics, known SARS-CoV-2 epidemiologic risk factors (e.g., an exposure in a nursing home), comorbid conditions, medications, Covid-19 related symptoms, and laboratory tests. Outcome of initial presentation to care (PTC), transfer to ICU, complications of Covid-19 disease, and 14-day clinical status were also extracted by manual review. PTC was defined as first contact with health care system with Covid-19 symptoms. The NIAID ordinal clinical outcome scale at 14 days from initial PTC was measured.^6^
 This report compares clinical and demographic characteristics, complications and 14-day outcomes for N=866 Covid-19 PCR positive patients hospitalized at MGH from March 11, 2020 – May 4, 2020.

We present results overall and stratified by self-identified race/ethnicity, with statistical testing comparing Hispanics and White/non-Hispanics. Tests of proportions (for binary outcomes) and t-tests (for numeric traits) were applied. All tests are two-sided and a p-value less than 0.05 was considered statistically meaningful. A multivariable logistic regression was fitted for a favorable outcome defined as hospital discharge or not requiring supplemental oxygen (ordinal score=1–3) at 14 days after PTC within the subset of individuals who were neither admitted to the ICU nor dead within this time frame. We evaluated the association of race/ethnicity with this outcome after adjusting for age, sex, body mass index (BMI), and smoking.

We additionally considered the degree of missing EHR data overall and by race/ethnicity among all Covid-19 confirmed cases hospitalized at MGH. Availability of BMI in the EHR over the past 3 years and number of visits per year were used as proxies for having prior comorbidity data that could inform Covid-19 risk factors and eligibility for clinical trials. The MGH Covid registry was approved by the Partners HealthCare Institutional Review Board (Protocol #2020P000829). Because this work was limited to chart review, individual consent was not obtained.

## Results

Overall, 43% of patients hospitalized with Covid-19 were women, median age was 60.4 [IQR=(48.2, 75)], 11.3% were Black/non-Hispanic and 35.2% were Hispanic (Table 1). Hispanic patients were younger than White/non-Hispanic patients [median age 51.1 years; IQR=(40.6, 61.6) versus 72 years; (58.0, 81.7) (p< 0.001)], less often ever smokers (23% vs 53%, p< 0.001) and more often had no known epidemiologic exposure (49% versus 35% p< 0.001) (Table 1). Hispanics were symptomatic for slightly longer before presenting (median 5 versus 3 days, p=0.039) and more commonly reported Covid-19 symptoms but were more likely to be sent home with self-quarantine rather than admitted to hospital at initial evaluation (29% versus 16%, p< 0.001). Documented co-morbidities were less common among Hispanics (e.g., 37% of Hispanics vs 62% of White/non-Hispanics had a history of hypertension, p< 0.001). Absolute lymphocyte count was higher and d-dimer lower on average among Hispanics at hospitalization (p< 0.001), while ferritin was higher among Hispanics compared to White/non-Hispanics (p=0.024).

**Table 1.** Risk factors and complications overall and by race/ethnicity^(a)^

|  | *Overall*<br>*(N=866)* | *White/*<br>*Non-Hispanic*<br>*(N=346)* | *Black/*<br>*Non-Hispanic*<br>*(N=98)* | *Hispanic*<br>*(N=305)* | *P-value for*<br>*test of White*<br>*vs. Hispanic* |
| --- | --- | --- | --- | --- | --- |
| **Presentation to Care (PTC)** |  |  |  |  |  |
| Female sex– count/total (%) | 375/866(0.43) | 147/346(0.42) | 51/98(0.52) | 138/305(0.45) | 0.529 |
| Age – Median (IQR) | 60.4 (48.2,75) | 72 (58,81.7) | 62.8 (52.8,72.3) | 51.1 (40.6,61.6) | <0.001 |
| Age >65 | 359/863(0.42) | 217/346(0.63) | 40/98(0.41) | 60/305(0.20) | <0.001 |
| Smoker (ever) | 329/861(0.38) | 184/345(0.53) | 38/98(0.39) | 70/301(0.23) | <0.001 |
| Body Mass Index | 29.1<br>(25.6,33.9) | 27.9<br>(24.8,33.4) | 28.8 (25,32.5) | 30.7 (26.6,34.8) | 0.030 |
| No known epidemiologic risk | 351/866(0.41) | 120/346(0.35) | 37/98(0.38) | 148/305(0.49) | <0.001 |
| Time from Symptoms to PTC | 4 (2,7) | 3 (1,7) | 3 (1,7) | 5 (3,8) | 0.039 |
| Fever | 567/866(0.65) | 191/346(0.55) | 55/98(0.56) | 231/305(0.76) | <0.001 |
| Cough without sputum | 451/866(0.52) | 164/346(0.47) | 45/98(0.46) | 176/305(0.58) | 0.011 |
| Dyspnea | 466/866(0.54) | 170/346(0.49) | 49/98(0.5) | 178/305(0.58) | 0.023 |
| Myalgia/Arthralgia | 332/866(0.38) | 100/346(0.29) | 29/98(0.3) | 151/305(0.5) | <0.001 |
| Diarrhea, nausea or vomiting | 286/866(0.33) | 100/346(0.29) | 29/98(0.3) | 121/305(0.4) | 0.005 |
| Statins | 362/866(0.42) | 187/346(0.54) | 33/98(0.34) | 93/305(0.3) | <0.001 |
| ACE Inhibitors or ARBs | 206/866(0.24) | 91/346(0.26) | 26/98(0.27) | 60/305(0.2) | 0.057 |
| Cardiac/metabolic disease history | 685/865(0.79) | 295/346(0.85) | 85/98(0.87) | 216/304(0.71) | <0.001 |
| Hypertension | 451/866(0.52) | 215/346(0.62) | 65/98(0.66) | 114/305(0.37) | <0.001 |
| Dyslipidemia | 324/866(0.37) | 150/346(0.43) | 36/98(0.37) | 93/305(0.3) | 0.001 |
| Pulmonary disease history | 270/862(0.31) | 145/343(0.42) | 30/98(0.31) | 64/304(0.21) | <0.001 |
| Diabetes (DM) type 2 | 299/866(0.35) | 118/346(0.34) | 42/98(0.43) | 96/305(0.31) | 0.529 |
| **Outcome at PTC** |  |  |  |  |  |
| Home with self-quarantine | 184/866(0.21) | 55/346(0.16) | 15/98(0.15) | 87/305(0.29) | <0.001 |
| Admitted to hospital | 528/866(0.61) | 229/346(0.66) | 63/98(0.64) | 176/305(0.58) | 0.032 |
| Admitted to ICU | 118/866(0.14) | 41/346(0.12) | 13/98(0.13) | 36/305(0.12) | 1.000 |
| **Admission labs<sup>(b)</sup>** |  |  |  |  |  |
| Absolute lymphocyte count < 0.80 | 295/808(0.37) | 158/326(0.48) | 25/93(0.27) | 71/281(0.25) | <0.001 |
| CRP > 100 | 294/781(0.38) | 104/314(0.33) | 36/92(0.39) | 101/272(0.37) | 0.353 |
| D-Dimer > 1000 | 371/753(0.49) | 164/297(0.55) | 56/87(0.64) | 107/267(0.40) | <0.001 |
| Ferritin > 500 | 400/787(0.51) | 142/315(0.45) | 41/92(0.45) | 150/274(0.55) | 0.024 |
| **Follow-up (14 day)** |  |  |  |  |  |
| In ICU or death during follow-up | 311/855(0.36) | 125/343(0.36) | 33/98(0.34) | 101/301(0.34) | 0.494 |
| ICU during follow-up<sup>(c)</sup> | 264/858(0.31) | 86/343(0.25) | 30/98(0.31) | 98/301(0.33) | 0.044 |
| Death during follow-up | 88/860(0.1) | 61/346(0.18) | 11/98(0.11) | 7/302(0.023) | <0.001 |
| ARDS | 199/863(0.23) | 69/346(0.20) | 18/98(0.18) | 72/302(0.24) | 0.269 |
| Liver dysfunction | 138/863(0.16) | 44/346(0.13) | 12/98(0.12) | 57/302(0.19) | 0.041 |
| Acute renal injury/failure | 175/863(0.20) | 79/346(0.23) | 24/98(0.24) | 46/302(0.15) | 0.019 |
| Ordinal Outcome Score<sup>(d)</sup> = 1-3 | 463/860(0.54) | 153/343(0.45) | 59/98(0.60) | 188/302(0.62) | <0.001 |
| Ordinal Outcome Score = 4-5 | 155/860(0.18) | 84/343(0.24) | 9/98(0.092) | 43/302(0.14) | 0.002 |
| Ordinal Outcome Score = 6-8 | 242/860(0.28) | 106/343(0.31) | 30/98(0.31) | 71/302(0.24) | 0.044 |

<sup>(a)</sup>Race/ethnicity breakdown: Additional patient categories not included in table: Asian (n=27), Other (n=22),
Missing (n=64), Native Hawaiian or other Pacific Islander (n=1), American Indian or Alaska Native (n=3). Results
are not reported separately for these categories due to the limited sample size. <sup>(b)</sup>Lab recorded within +/-3 days of
hospital admission. <sup>(c)</sup> 47 patients died without being admitted to the ICU (39 White, 3 Hispanic, 3 Black, 1 Asian, 1
Missing). <sup>(d)</sup>Score: 8 – death; 7 – hospitalized, on invasive mechanical ventilation or ECMO; 6 – hospitalized, on
non-invasive ventilation or high flow oxygen; 5 – hospitalized requiring supplemental oxygen; 4 – hospitalized, not
requiring supplemental oxygen, requiring ongoing medical care; 3 – hospitalized, not requiring oxygen, no longer
requiring ongoing medical care; 2 – not hospitalized, limitation on activities and/or requiring home oxygen; 1 – not
hospitalized, no limitations on activities

During the study period, ICU bed capacity was sufficient. In the 14 days following presentation to care, 36% of patients went to the ICU or died prior to going to the ICU. Although a comparable proportion of Hispanics and White/non-Hispanics received ICU care or died (34% versus 36%) and experienced ARDS (24% versus 20%), a larger proportion of Hispanics experienced liver dysfunction (19% vs 13%, p=0.041) and a smaller proportion had acute renal injury/failure (15% versus 23%, p=0.019). While there were few deaths by 14 days from PTC (N=88, 10%), a smaller proportion of Hispanic patients had died compared to White/non-Hispanic patients (2% versus 18%, p< 0.001). A larger proportion of Hispanics had the best outcome score at 14 days (score 1–3, 62% versus 45%, p< 0.001). Among patients neither admitted to the ICU nor dead within 14 days of PTC, Hispanics were significantly more likely to have the best outcome score (OR=1.99, p=0.011) compared to White/non-Hispanics after controlling for age, sex, BMI and smoking status.

Many hospitalized patients, and disproportionately Hispanic patients, were new to our health system, as measured by availability of BMI in the EHR. A smaller proportion of Hispanics than White/non-Hispanics had a BMI measurement in the EHR in 2019 (37% versus 62%) and for the three-year period 2017–2019 (16% versus 40%). The median number of months since first contact was 20.5 for Hispanics and 69.0 for White/non-Hispanics and Hispanic patients had fewer visits/year among years with available data (1.5 versus 2.3).

## Discussion

Among patients hospitalized for Covid-19 at MGH, 35% were Hispanic, although Hispanic patients comprised 8% of inpatient discharges at MGH in 2017.^7^
 Hispanic patients were younger with fewer comorbidities associated with severe Covid-19 disease, yet had comparable rates of ICU care or death as White/non-Hispanics.^8^
 Despite similarly severe clinical courses, a greater proportion of Hispanics recovered by 14 days after PTC and fewer died. While older age is associated with hospitalization for Covid-19 infection in the US, hospitalized Hispanic patients in our sample were substantially younger than their White/non-Hispanic counterparts.^9^
 Older patients may have been more likely to have “do not intubate” directives and thus be less likely to transfer to the ICU; we therefore used the composite severe outcome of ICU or death to minimize confounding. Hispanics had fewer comorbidities and were less likely to experience acute kidney injury and these may explain the better clinical outcomes that we observed. However, longer term follow-up and judicious consideration of additional information, including age^10^
 and socioeconomic factors^11^
 are needed to better evaluate the significance of these findings.

A strength of this study was the use of manual chart review for identifying comorbid conditions. We found that fewer Hispanic patients had data in our EHR in the three years preceding the appearance of SARS-CoV-2. Underreporting of pre-existing comorbidities in observational studies, from limited EHR data or under-diagnosis, can bias the relationship between comorbidities and clinical outcomes differentially across marginalized groups.^12^
 This must be considered when studies rely on EHR data for assessing Covid-19 risk factors or clinical outcomes. A limitation of this study is that due to sample size, we were unable to meaningfully assess differences among other racial and ethnic groups.

Hispanics are disproportionately impacted by Covid-19 and have a severe disease despite younger age and fewer documented comorbidities. This underscores the urgent need to report disaggregated outcomes to better understand Covid-19 among racial/ethnic groups. The design and practice of clinical trials for treatments and vaccines can benefit from a deeper understanding of the profound heterogeneity of the populations Covid-19 impacts. As measures are taken to fortify public health infrastructure in the United States, consideration of potential racial/ethnic differences in presentation and access to care as well as in clinical outcomes is essential to the reduction of health disparities.

## Data Availability

Data are available by request to with appropriate permissions;.

## Acknowledgements

The authors gratefully acknowledge the dedicated manual chart reviewers and experienced data managers for their methodological expertise, time and effort.

